# Association of Cannabis Legalization with Cannabis Positive Drug Screening in US Veterans

**DOI:** 10.1101/2023.12.08.23299731

**Authors:** David S. Fink, Hillary Samples, Carol A. Malte, Mark Olfson, Melanie M. Wall, Daniel M. Alschuler, Andrew J. Saxon, Deborah S. Hasin

## Abstract

**Background:** Although cannabis legalization is associated with increases in self-report cannabis use, biological measures of cannabis use are needed to address potential bias introduced by improved self-reporting of cannabis use in states enacting medical cannabis laws (MCL) and recreational cannabis laws (RCL).

**Objective:** Quantify the role of MCL and RCL enactment in cannabis positive urine drug screen (UDS) prevalence among Veterans Health Administration (VHA) emergency department (ED) patients from 2008 to 2019.

**Design:** Staggered-adoption difference-in-difference analysis were used to estimate the role of MCL and RCL in cannabis positive UDS data, fitting adjusted linear binomial regression models to estimate the association between MCL and RCL enactment and prevalence of cannabis positive UDS.

**Participants:** VHA enrolled veterans aged 18-75 years with ≥1 ED visit in a given year from 2008 to 2019.

**Main Measures:** Receipt of ≥1 cannabis positive UDS during an ED visit were analyzed.

**Key Results:** From 2008 to 2019, adjusted cannabis positive UDS prevalences increased from 16.4% to 25.6% in states with no cannabis law, 16.6% to 27.6% in MCL-only enacting states, and 18.2% to 33.8% in RCL-enacting states. MCL-only and MCL/RCL enactment was associated with a 0.8% (95% CI, 0.4-1.0) and 2.9% (95% CI, 2.5-3.3) absolute increase in cannabis positive UDS, respectively. Significant effect sizes were found for MCL and RCL, such that 7.0% and 18.5% of the total increase in cannabis positive UDS prevalence in MCL-only and RCL states could be attributed to MCLs and RCLs.

**Conclusions:** In this study of VHA ED patients, MCL and RCL enactment played a significant role in the overall increases in cannabis positive UDS. The increase in a biological measure of cannabis use reduces concerns that previously documented increases in self-reported cannabis use from surveys are due to changes in patient willingness to report use as it becomes more legal.

## Introduction

As of November 2023, 39 states have legalized medical cannabis use and 24 states and Washington D.C. have legalized recreational use by adults. Based on self-report data from national surveys, such legalization is associated with increased rates of cannabis use.^1,2^ However, the validity of self-reported drug use has long been controversial due to concerns that individuals may under-report drug use for various reasons (e.g., perceived stigma; fear of adverse social, legal or treatment consequences), thereby biasing study results. In clinical trials, biological indicators of drug use, e.g., urine toxicology testing, are often used as outcome measures to mitigate such potential bias.^3,4^ Yet while the challenges and expense of such measures are generally feasible in clinical studies, they are generally unfeasible in large-scale observational studies of issues such as time trends in drug use. Therefore, much of the extant literature on time trends is based on self-reported measures of drug use that are vulnerable to reporting biases.

Even when survey confidentiality is promised, self-reports of drug use can be biased by contextual factors that can change over time as the legal and social environment surrounding the use of a substance evolves. Studies of time trends in self-reported cannabis use may be particularly vulnerable to changing contextual bias due to the marked changes in environmental factors (e.g., legalization; acceptability) over the last 25 years. Therefore, information on time trends in the prevalence of cannabis utilizing biological measures, if available, would represent a substantial methodological advance. We identified one such source, the electronic medical record urine toxicology data available across years from the Veterans Health Administration (VHA). We leveraged these data to examine the relationship of MCL and RCL enactment to cannabis use as indicated by cannabis-positive urine drug screens (UDS) in VHA emergency department patients.

## Methods

Data from Veterans Health Administration (VHA) patients aged 18-75 years with ≥1 emergency department (ED) visit in a given year between 2008 to 2019 were analyzed, excluding those who received hospice/palliative care or resided outside the U.S. Data from ED visits were analyzed because rates of UDS for cannabis were steady over the study period (10.0% in 2008, 11.9% in 2019). With these data, we created 12 yearly datasets from. Primary exposures were state-year variables indicating state enactment of MCLs and RCLs, i.e., that the law was operational, and residents could rely on its legal protection. Patient state of residence was indicated by location of last healthcare encounter for each year. States were categorized each year as no cannabis laws (no CLs), MCL-only, and MCL/RCL (no states have enacted RCL without previously enacting MCL). All dates were provided by RAND-USC Opioid Policy Tools and Information Center (OPTIC) marijuana policy data.^5^ The main outcome was ≥1 UDS positive for Δ-9-tetrahydrocannabinol (THC) metabolite during an ED visit in a given year.^6^ To estimate the role of MCL and RCL enactment in the national increases in cannabis positive UDS prevalence using all yearly information from 2008 to 2019, a staggered adoption difference-in-difference (DiD) model was used.^7^ The DiD estimates for MCL-only and MCL/RCL associations were obtained from a linear binomial regression model with fixed effects for state, categorical year, time-varying law status, individual-level covariates (age, sex, race/ethnicity), and time-varying state-level covariates (age, sex, race/ethnicity, income, poverty, unemployment). To illustrate the magnitude of the DiD estimate compared with the overall increase in cannabis-positive UDS prevalence (i.e., the amount of change that could be attributed to the laws), DiD estimates were divided by the absolute changes between 2008 and 2019 in the states with the respective laws by 2019.^8^ The study was approved by Institutional Review Boards at New York State Psychiatric Institute, VHA Puget Sound and VHA New York Harbor Healthcare Systems.

## Results

The ED samples ranged from 790,754 to 1,020,865 patients per year., among patients that received UDS in 2008 and 2019, 16.42% and 27.8% were positive for cannabis, respectively. **Figure 1** and **Table 1** show 2008 to 2019 cannabis positive UDS prevalence trends (weighted mean estimates) within no-CL, MCL-only, and MCL/RCL states defined by their 2019 status. Cannabis-positive UDS prevalence increased in all 3 groups of states: from 16.4% to 25.6% in no-CL states (9.3% absolute increase), from 16.6% to 27.6% in MCL-only states (11.0% absolute increase), and from 18.2% to 33.8% in MCL/RCL states (15.7% absolute increase).

**Table 1.** Adjusted positive urine drug screening (UDS) prevalence^*^ in VHA patients treated in the emergency department in 2008 and 2019, by enacted state law status, absolute change over time, and DiD estimates.

| Type of State | Cannabis positive UDS prevalence, %<sup>†</sup> |  | Absolute Change % | Model-based DiD law result, % (95%CI)<sup>□</sup> | P value | % of absolute change attributable to law<sup>‡</sup> |
| --- | --- | --- | --- | --- | --- | --- |
|  | 2008 | 2019 |  |  |  |  |
| No-CL (17 states by 2019) | 16.4 | 25.6 | 9.3 | Reference |  |  |
| MCL-Only (22 states by 2019) | 16.6 | 27.6 | 11.0<sup>‡</sup> | 0.8 (0.5, 1.0) | < .001 | 7.0% |
| MCL/RCL (12 states by 2019) | 18.2 | 33.8 | 15.7<sup>§</sup> | 2.9 (2.5, 3.3) | < .001 | 18.5% |

Abbreviations: CL, cannabis law; MCL, medical cannabis law; RCL, recreational cannabis law; VHA, veteran health administration; UDS, urine drug screening
\*Adjusted for categorical age, sex and race/ethnicity, yearly state-level median income, and yearly state rates of male individual, Hispanic individuals, non-Hispanic Black individuals, non-Hispanic White individuals, those in poverty category, those 18 and older, and those who are unemployed. <sup>†</sup>Adjusted for continuous age, sex and race/ethnicity, yearly state-level median income, and yearly state rates of male individual, Hispanic individuals, non-Hispanic Black individuals, non-Hispanic White individuals, those in poverty category, those 18 and older, and those who are unemployed. <sup>‡</sup>Value used as denominator to determine the % of overall increase attributable to MCL enactment based on difference-in-difference estimates of MCL effect sizes. <sup>§</sup>Value used as denominator to determine the % of overall increase attributable to RCL enactment based on difference-in-difference estimates of RCL effect sizes. <sup>□</sup>Staggered-Adoption Difference-in-Difference (DID) model<sup>7</sup>, adjusted for continuous age, sex and race/ethnicity. <sup>‡</sup>DiD estimate divided by absolute change across period

**Figure 1.**
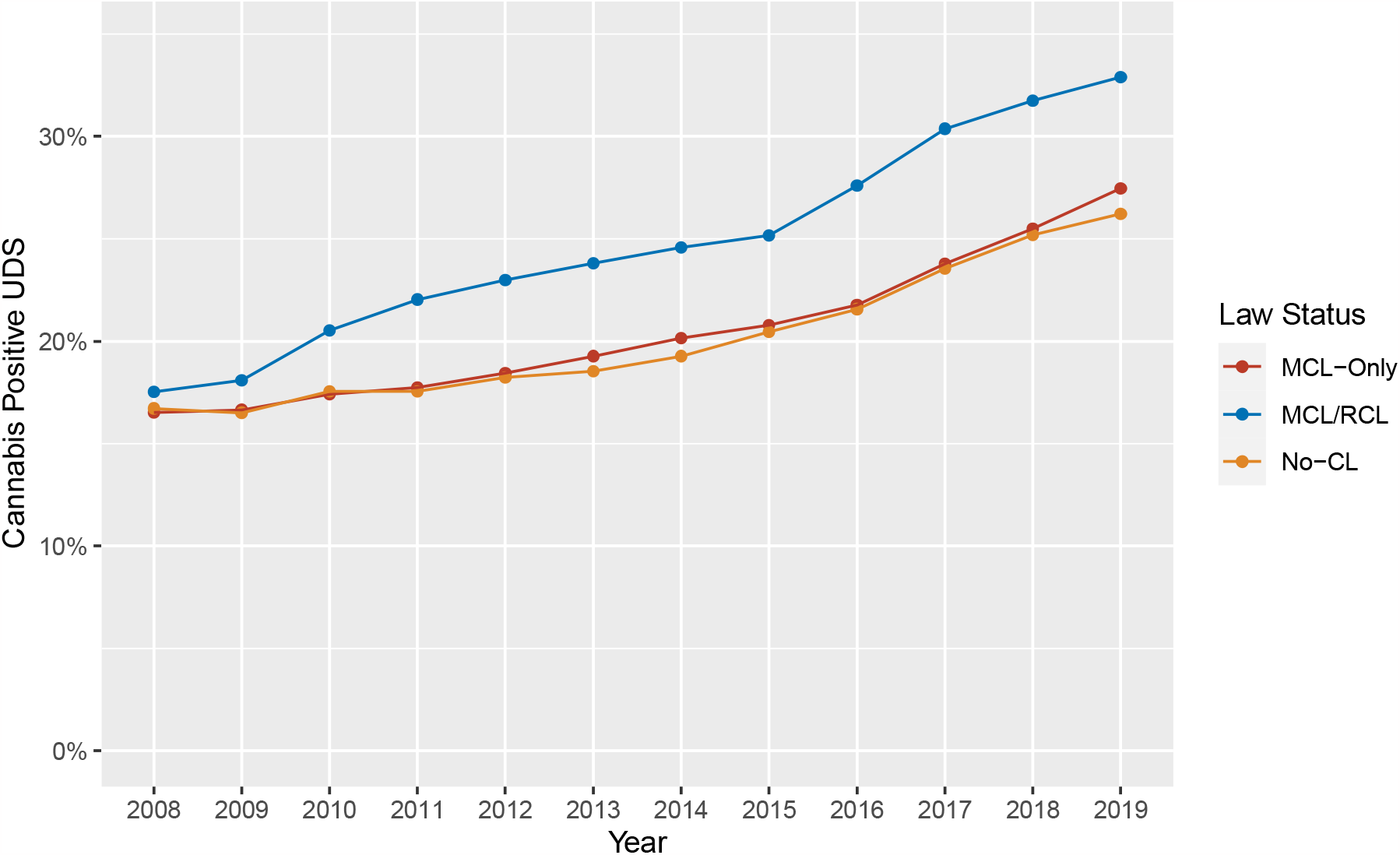
Trends in cannabis positive urine drug screens (UDS) (weighted mean estimates adjusted for age, sex, and race/ethnicity) from 2008 to 2019, aggregated within the three groups of states defined by their status at the end of 2019 Weighted mean prevalence estimates adjusted for age, sex, race and ethnicity and time-varying state covariates are reported. Error bars indicate 95%CIs; CIs are very small because of the large sample sizes. CL indicates cannabis law; MCL, medical cannabis law, RCL, recreational cannabis law, UDS, urine drug screening

The DiD estimate of the prevalence increase in cannabis-positive UDS due to MCL enactment was 0.8% (95% CI, 0.5%-1.0%; **Table 1**). Relative to the absolute change in MCL-only states by 2019 (11.0%; **Table 1**), this estimate indicates that 7.0% of the increase in cannabis-positive UDS could be attributed to MCL-only enactment. The DiD estimate of the cannabis-positive UDS prevalence increase due to MCL/RCL enactment was 2.9% (95% CI, 2.5%, 3.3%). Relative to the absolute change in MCL/RCL states by 2019 (15.7%; **Table 1**), 18.5% of the increase in RCL states could be attributed to RCL enactment.

## Discussion

Using urine drug screen (UDS) data from VHA emergency department patients between 2008 and 2019, we found that cannabis-positive UDS prevalence increased in no-CL, MCL-only, and MCL/RCL states. While no-CL and MCL-only states followed a similar trajectory over time, MCL/RCL states had the greatest relative increase in cannabis-positive UDS. Difference-in-differences models that adjusted for contemporaneous trends before and after MCL or RCL enactment suggested that cannabis-positive UDS prevalence in ED patients increased after MCL enactment by 0.77% more than would have occurred in the absence of MCLs, and by an additional 2.89% after RCL enactment than would have occurred in the absence of RCLs. These increases represent the estimated effects on cannabis-positive UDS prevalence specifically associated with enactment of the laws and suggest that of the total 2008 to 2019 increase in cannabis-positive UDS prevalence in this national patient population, MCL and RCL accounted for 7.0% and 18.5%, respectively.

Urine drug screens to test for substance use represents an important additional measure of cannabis use for understanding changes in cannabis use following MCL and RCL enactment. Previous national studies of MCL and RCL found post-enactment increases in the prevalence of self-reported use ^2,9,10^ and also in clinical diagnosis for cannabis use disorder (CUD) diagnoses in VHA patients, diagnoses that cannot be made without self-reported cannabis use.^8^ Cannabis legalization could reduce concerns about disclosing cannabis use in surveys and clinical settings, potentially leading to artifactual findings on prevalence increases and MCL and RCL effects. An earlier VHA study showed increases in CUD diagnoses associated with MCL and RCL enactment from 2005 to 2019 among the overall VHA patient population^8^. Since our UDS data do not rely on self-report or clinical attention to cannabis use disorder symptoms, the present study provides an important new source of data to understand the effects of MCL and RCL policies. The results reduce concerns that the previous findings on self-reported cannabis use and CUD diagnoses following cannabis legalization are not due to post-policy changes in patient willingness to report use as it becomes legalized.

This study has limitations. Findings may not generalize to veterans enrolled in other public or private insurance programs, to uninsured veterans or nonveteran populations.^11^ Also, we were unable to differentiate between cannabinoid use for medical or recreational purposes. Nevertheless, our results identified increases in cannabis use after MCL and RCL enactment, supporting a role of the policy environment in contributing to the widespread increase in cannabis use among US adults. Finally, RCL and MCL policies may change ED ordering of UDS and patients’ willingness to agree to UDS testing. However, the consistent level of UDS testing over the study period mitigates this concern.

Our study extends findings from the extant literature by using a biological measure of cannabis use to analyze the association between MCLs and RCLs on cannabis use. Although prior research consistently shows that the prevalence of self-reported cannabis use increases after states enact MCLs^2,12,13^ and RCL,^14^ self-report measures may be subject to recall and social desirability biases. Urine drug screen data provide a novel and important source of information on the effects of state legalization of medical and recreational cannabis use. While increases in social acceptability of use and policies allowing use may attenuate the effects of social desirability bias over time, changes in social desirability bias are unlikely to affect UDS data collected in ED patients, which are generally done to target drugs other than cannabis. Therefore, clinicians in states with legal cannabis should pay especially close attention to monitoring for cannabis and consideration of whether problems exist in patients who use cannabis.

## Data Availability

The data are not available.

## Author Contributions

Dr. Fink had full access to all of the data in the study and takes responsibility for the integrity of the data and the accuracy of the data analysis.

*Concept and design:* Hasin, Wall, Olfson, Fink, Saxon.

*Acquisition, analysis, and interpretation of data:* Fink, Malte, Alschuler, Wall, Saxon, Hasin.

*Drafting of the manuscript:* Fink, Samples

*Critical revision of the manuscript for important intellectual content:* Fink, Samples, Malte, Alschuler, Wall, Saxon, Hasin

*Statistical analysis:* Fink

*Obtained funding:* Fink, Hasin.

*Administrative, technical, or material support:* Fink, Malte, Alschuler, Wall, Saxon, Hasin.

*Supervision:* Wall, Saxon, Hasin.

## Data available

No

## References

1. Cerdá M, Mauro C, Hamilton A, et al. Association Between Recreational Marijuana Legalization in the United States and Changes in Marijuana Use and Cannabis Use Disorder From 2008 to 2016. JAMA psychiatry. 2020;77(2):165–171.

2. Martins SS, Mauro CM, Santaella-Tenorio J, et al. State-level medical marijuana laws, marijuana use and perceived availability of marijuana among the general U.S. population. Drug and alcohol dependence. 2016;169:26–32.

3. Shulman M, Weiss R, Rotrosen J, Novo P, Costello E, Nunes EV. Prior National Drug Abuse Treatment Clinical Trials Network (CTN) opioid use disorder trials as background and rationale for NIDA CTN-0100 “optimizing retention, duration and discontinuation strategies for opioid use disorder pharmacotherapy (RDD)”. Addiction Science & Clinical Practice. 2021;16(1):15.

4. Incze MA. Reassessing the Role of Routine Urine Drug Screening in Opioid Use Disorder Treatment. JAMA internal medicine. 2021;181(10):1282–1283.

5. RAND Corporation. OPTIC-Vetted Policy Data Sets. RAND Corporation. https://www.rand.org/health-care/centers/optic/resources/datasets.html. Published 2022. Accessed.

6. Fink DS, Malte C, Cerdá M, et al. Trends in Cannabis-positive Urine Toxicology Test Results: US Veterans Health Administration Emergency Department Patients, 2008 to 2019. Journal of addiction medicine. 9900:10.1097/ADM.0000000000001197.

7. Athey S, Imbens GW. Design-based analysis in Difference-In-Differences settings with staggered adoption. Journal of Econometrics. 2022;226(1):62–79.

8. Hasin DS, Wall MM, Choi CJ, et al. State Cannabis Legalization and Cannabis Use Disorder in the US Veterans Health Administration, 2005 to 2019. JAMA psychiatry. 2023;80(4):380–388.

9. Weinberger AH, Wyka K, Kim JH, et al. A difference-in-difference approach to examining the impact of cannabis legalization on disparities in the use of cigarettes and cannabis in the United States, 2004-17. Addiction. 2022;117(6):1768–1777.

10. Hasin DS, Sarvet AL, Cerdá M, et al. US Adult Illicit Cannabis Use, Cannabis Use Disorder, and Medical Marijuana Laws: 1991-1992 to 2012-2013. JAMA psychiatry. 2017;74(6):579–588.

11. Fink DS, Stohl M, Mannes ZL, et al. Comparing mental and physical health of U.S. veterans by VA healthcare use: implications for generalizability of research in the VA electronic health records. BMC health services research. 2022;22(1):1500.

12. Wen H, Hockenberry JM, Cummings JR. The effect of medical marijuana laws on adolescent and adult use of marijuana, alcohol, and other substances. Journal of Health Economics. 2015;42:64–80.

13. Hasin DS, Sarvet AL, Cerda M, et al. US Adult Illicit Cannabis Use, Cannabis Use Disorder, and Medical Marijuana Laws: 1991-1992 to 2012-2013. JAMA psychiatry. 2017;74(6):579–588.

14. Cerdá M, Mauro C, Hamilton A, et al. Association Between Recreational Marijuana Legalization in the United States and Changes in Marijuana Use and Cannabis Use Disorder From 2008 to 2016. JAMA psychiatry. 2019;77(2):165–171.

